# Automated Detection of Lacunes in Brain MR Images Using SAM with Robust Prompts via Self-Distillation and Anatomy-Informed Priors

**DOI:** 10.1101/2025.07.09.25331177

**Authors:** Pon Deepika, Gouri Shanker, Ramanujam Narayanan, Vaanathi Sundaresan

**Affiliations:** Department of Computational and Data Sciences, Indian Institute of Science, Bengaluru, Karnataka 560012, India

**Keywords:** Cerebral small vessel disease, Lacune detection, Segment Anything Model, Region-aware thresholds, Contrastive learning, Self-distillation, Simulated lesions

## Abstract

Lacunes, which are small fluid-filled cavities in the brain, are signs of cerebral small vessel disease and have been clinically associated with various neurodegenerative and cerebrovascular diseases. Hence, accurate detection of lacunes is crucial and is one of the initial steps for the precise diagnosis of these diseases. However, developing a robust and consistently reliable method for detecting lacunes is challenging because of the heterogeneity in their appearance, contrast, shape, and size. To address the above challenges, in this study, we propose a lacune detection method using the Segment Anything Model (SAM), guided by point prompts from a candidate prompt generator. The prompt generator initially detects potential lacunes with a high sensitivity using a composite loss function. The SAM model selects true lacunes by delineating their characteristics from mimics such as the sulcus and enlarged perivascular spaces, imitating the clinicians’ strategy of examining the potential lacunes along all three axes. False positives were further reduced by adaptive thresholds based on the region-wise prevalence of lacunes. We evaluated our method on two diverse, multi-centric MRI datasets, VALDO and ISLES, comprising only FLAIR sequences. Despite diverse imaging conditions and significant variations in slice thickness (0.5–6 mm), our method achieved sensitivities of 84% and 92%, with average false positive rates of 0.05 and 0.06 per slice in ISLES and VALDO datasets respectively. The proposed method outperformed the state-of-the-art methods, demonstrating its effectiveness in lacune detection and quantification.

## 1 Introduction

Lacunes of presumed vascular origin appear as round or ovoid subcortical fluid-filled cavities, between 3 and approximately 15 mm in diameter, as defined by the STandards for ReportIng Vascular changes on nEuroimaging (STRIVE) criteria (Wardlaw et al., 2013; Duering et al., 2023). Although lacunes are frequently observed in older adults and are often asymptomatic (Wardlaw, 2008), they have been associated with stroke, dementia, cognitive decline, and gait impairments (Jokinen et al., 2011; Vermeer et al., 2003; Chen et al., 2020). Accurate detection and quantification of lacunes are essential for patient risk stratification, enabling the precise diagnosis of cerebrovascular and neurodegenerative diseases and ensuring timely treatment. In FLAIR sequences, lacunes generally appear as round or ovoid hypointense regions, similar to the CSF (indicating fluid-filled cavities), surrounded by an irregular hyperintense rim (Duering et al., 2023). However, this characteristic rim can also be partially or completely absent, depending on various factors, such as the extent of gliosis, the age of the lesion, and its location (Duering et al., 2023, 2013). This contributes to the significant diversity in the appearance of lacunes, as shown in Figure 1, making visual assessment a challenging and time-consuming task for radiologists. The accurate detection of lacunes is further complicated by the presence of mimics, including anatomical structures (e.g., sulci, cerebral ventricle) and other imaging biomarkers of cerebral small vessel disease (CSVD), such as enlarged perivascular spaces (EPVS) (Duering et al., 2023; Uchiyama et al., 2007a). In particular, EPVS are fluid-filled spaces that typically follow the course of the vessel, exhibiting a signal similar to CSF, and appear round or ovoid when imaged perpendicular to the vessel’s course. This makes them indistinguishable from lacunes when located within white matter hyperintensities and exceeds 3 mm in size (Duering et al., 2023).

**Figure 1:**
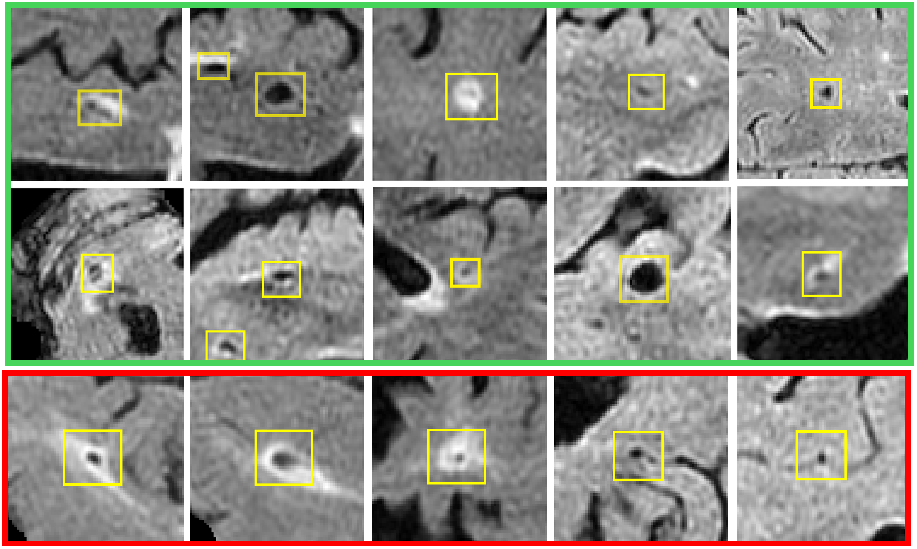
Heterogeneity in the appearance of lacunes on fluid-attenuated inversion recovery (FLAIR) sequence. The first two rows show the diversity in lacunes (indicated by green box), while the third row illustrates non-lacune mimics (indicated by red box).

Given the inherent complexities in lacune detection, manual lacune labelling is highly time intensive, expensive for larger datasets, and requires a high level of expertise. Additionally, labelling may be prone to high inter-rater variability. Hence, there is a growing need for computer-aided methods to detect lacunes efficiently. While a few semi-automated and automated methods have been proposed, as detailed in Section 1.1, existing approaches often require human intervention (in the case of semi-supervised methods), provide suboptimal performance due to false detections of mimics, or lack generalisability across multiple resolutions. To address these limitations, we propose a novel fully automated framework for effectively identifying lacunes in brain MRI scans across different image resolutions. The proposed method involves a modular approach comprising lacune candidate detection, identification of lacunes from mimics using representative prompts, and further reduction of false positives using adaptive thresholding. The key contributions of our method include:

1. Developing a Candidate Prompt Generator (CPG), a deep learning module that employs a combination of loss functions to differentiate between tissue characteristics and generate precise lacune candidate prompts to guide the SAM detection process.
2. Leveraging the Segment Anything Model (SAM) to identify true lacunes by analysing the candidate regions along all three anatomical planes, replicating the decision making process of clinical experts.
3. Utilising the prior knowledge of regional lacune distribution at population-level to determine adaptive thresholds for false positive reduction, ensuring high detection accuracy.
4. Implementing a self-distillation approach to significantly enhance lacune detection performance on a smaller dataset.

Lacune identification often requires a method that functions robustly at various image resolutions. Hence, we evaluated the effectiveness of our method using two datasets consisting of MRI volumes from four different centres with slice thicknesses ranging from 0.5 to 6 mm. We also conducted ablation studies to evaluate the influence of different components of loss functions and the effect of using simulated lesions in the training data on effective candidate detection. In addition, we performed ablation studies to examine the contribution of different modules to the detection of lacunes. We also compared our results with existing state-of-the-art methods for lacune detection.

### 1.1 Related Work

Several semi-automated and automated methods have been proposed to address the challenges associated with lacune detection. For example, Al-Masni *et al*. (Al-Masni et al., 2021) proposed a semi-supervised method consisting of a candidate discriminator based on a 3D multiscale residual network to distinguish true lacunes from lacune mimics identified by neuroradiologists. However, manual intervention was required for candidate detection, which was challenging for a wide range of mimics.

Early work on automated methods for lacune detection by Yokoyama *et al*. (Yokoyama et al., 2007) involved a comprehensive method that combined multiphase binarisation, morphological operations, and feature-based classification. Given the challenges of the accurate detection of lacunes, researchers have often adopted multi-stage approaches consisting of potential candidate identification, followed by systematic refinement. In this context, Uchiyama *et al*. investigated various techniques for false positive reduction by employing rule-based algorithms (Uchiyama et al., 2007a), support vector machines (Uchiyama et al., 2012), neural network classifiers (Uchiyama et al., 2007b, 2008), and eigen-template matching (Uchiyama et al., 2015). Furthermore, using conventional methods, Wang *et al*. (Wang et al., 2012) introduced a multistage framework for segmenting white matter hyperintensities, cortical infarcts, and lacunar infarcts, relying on a rule-based approach driven by absolute voxel intensities. However, these conventional methods face significant challenges in learning robust representations of lacunes, particularly because lacunes are small and their appearance varies with slice thickness, imaging protocols, and the age of the lesion, as evidenced by the high false positive rates of these studies (e.g., 0.47 false positives per slice reported in

With advancements in computational capabilities, various deep learning-based methods have recently been explored for detecting lacunes, demonstrating promising performance compared to conventional techniques (Sudre et al., 2019; Al-Masni et al., 2021; Ghafoorian et al., 2017). In particular, Ghafoorian *et al*. (Ghafoorian et al., 2017) developed a two-stage deep learning approach to detect potential lacunes, where the initial extraction of candidates was performed using a patch-based 2D CNN segmentation model. To address false positives from the candidate extractor and differentiate lacunes from mimics, several studies, including Ghafoorian et al. (Ghafoorian et al., 2017) have investigated multi-scale 3D patch analysis (Ghafoorian et al., 2017; Al-Masni et al., 2021). However, their evaluation was limited to uniform resolution volumes, leaving the effectiveness of the method on multi-resolution data unexplored. Other deep learning-based frameworks explored for lacune detection included U-Net (Duan et al., 2020) and adaptations of the RCNN network for 3D data (Sudre et al., 2019). Despite these advancements, the accurate detection of lacunes remains a significant challenge owing to their diverse appearance, size, and presence of mimics. These factors frequently contribute to high false positive rates, with the lowest reported rate being 0.13 per slice (Ghafoorian et al., 2017).

The VAscular Lesions DetectiOn and Segmentation Challenge (VALDO 2021) (Sudre et al., 2024), held in conjunction with MICCAI 2021, the largest platform organised so far for accurate lacune detection, invited research groups to develop automated solutions for detecting and segmenting subtle signs of CSVD, including lacunes. Various methods have been developed for VALDO 2021, including versions of UNet, MaskRCNN, and MaskRetinanet, with different loss functions, such as Dice, blob, binary cross-entropy, focal, or mean absolute error. The top-ranking team employed a 3D UNet model with Dice loss based on the nnUNet framework. Their method achieved a median F1 score of 28.57, indicating the scope for improvement with the need for more advanced deep learning techniques.

A major challenge in the development of supervised automated methods for lacune detection is the time-consuming process of annotating small lesions. This is further complicated by the limited availability of training data in various datasets, which affects model generalisation. Recent advances in foundation models, such as the Segment Anything Model (SAM) (Kirillov et al., 2023) and its variants, have been explored to address challenges related to limited data and model generalisability by harnessing their zero-shot segmentation capabilities. SAM has shown promising performance in segmenting various lesions and organs using multiple imaging modalities, including CT, MRI, ultrasound, and PET, using appropriate prompts without requiring additional training (Mazurowski et al., 2023). However, the utility of SAM in detecting smaller lesions such as lacunes remains largely unexplored. Similarly, techniques, such as adversarial learning and feature alignment strategies, have been widely explored (Singhal et al., 2023) to overcome training data scarcity and shifts across datasets. Among these, knowledge distillation has provided improved performance for various tasks, including multi-organ segmentation (Kim et al., 2024), vertebrae detection (Serrador et al., 2024), brain tumour analysis (Choi et al., 2023), white matter hyperintensity segmentation (Orbes-Arteainst et al., 2019), microbleed detection (Sundaresan et al., 2023), and retinal disease classification (Paluru et al., 2023), illustrating its effectiveness in improving model generalisability, even with limited target training data. Hence, given the challenges in lacune detection, as highlighted by prior works (Al-Masni et al., 2021; Ghafoorian et al., 2017), there is a need for specialised modular frameworks using foundation models such as SAM with accurate prompts. Additionally, it would be desirable to address challenges such as wide appearance heterogeneity, multi-centre variability, and limited data availability, using techniques such as knowledge distillation, for accurate and generalisable detection of lacunes.

## 2 Materials and methods

In the proposed lacune detection workflow, we initially preprocessed the MRI volumes (Section 2.1) and identified potential lacune candidates using the candidate prompt generator (CPG) (Section 2.2). The aim of the CPG module was to provide precise prompts for SAM. We later performed a systematic detection of true lacunes among the candidates from the CPG module by analysing these prompts across all three anatomical planes using SAM (Section 2.3). Finally, we further improved the lacune-detection accuracy using an adaptive region-aware thresholding (ART) module (Section 2.4). Figure 2 illustrates the workflow of the proposed lacunar detection method.

**Figure 2:**
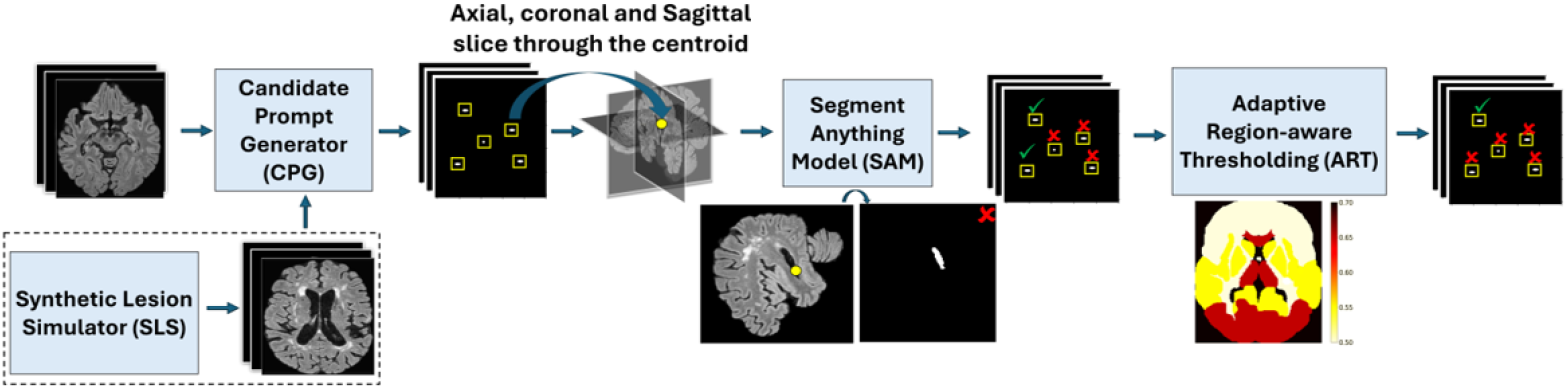
Workflow of the proposed lacune detection method. The initial lacune candidates that are detected by Candidate Prompt Generator (CPG) are used to extract point prompts for Segment Anything Model (SAM) model for lacune detection from mimics. The Adaptive Region-aware thresholding (ART) module is applied on the SAM output to further reduce the false detections. The CPG module is trained using real-world lacunes and synthetic lacune lesions generated by the Synthetic Lesion Simulator (SLS) module (used only for CPG training) as indicated by the dotted box.

### 2.1 Data preprocessing

We skull-stripped the FLAIR volumes using FSL BET (Smith, 2002), and subsequently corrected them for bias field inhomogeneities using FSL FAST (Zhang et al., 2001). Finally, we cropped the images closer to the edges of the brain to create a tight field of view.

### 2.2 Candidate prompt generator

As a first step towards accurate detection of lacunes, the aim of the CPG module was to identify potential lacune candidates on brain MRI volumes with high sensitivity, and later remove the false positives in the subsequent steps. In the CPG module, we developed a deep learning model that focused on segmenting lacunes from 2D axial slices. We chose to use a 2D model because of the high interslice variation of lacunes across various datasets. Axial slices were used for the 2D model because the spatial resolution was generally optimal in axial slices across the datasets for detecting lacunes. All 2D axial slices were z-score normalised for the CPG.

We selected the *U*^2^Net (Qin et al., 2020) architecture in the CPG module to segment the initial lacune candidates. *U*^2^Net uses Residual U-Blocks (described in (Qin et al., 2020)) to efficiently capture features at multiple scales. This design enabled the model to learn more complex patterns of lacunes, as shown in Figure 1. *U*^2^Net provided better sensitivity for the detection of lacune candidates than the other architectures, as described in Section 4.1.

We utilised a composite loss function, that integrated Tversky (Salehi et al., 2017), focal (Ross and Dollár, 2017), and pixel-wise contrastive losses (Khosla et al., 2020) for precise pixel-wise learning of lacune characteristics. First, to maximise recall by detecting as many true lacunes as possible, we used the Tversky loss (*L*_*tver*_) given in Equation 1 with a high penalty for false negatives.

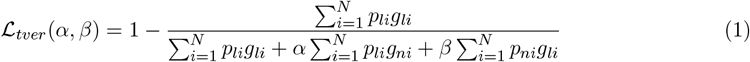

where *p*_*ni*_ and *p*_*li*_ are the predicted probabilities of pixel *i* being a non-lacune and lacune, respectively. The indicator variables *g*_*ni*_ and *g*_*li*_ are 1 for non-lacune and lacune pixels, respectively, and 0 otherwise. Second, to overcome the class imbalance between lacunes and the background in lacune candidate segmentation, we added focal loss (*L*_*foc*_) (Eqn. 2) to help the model focus more on challenging borderline pixels, further enhancing performance.

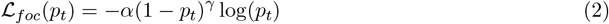

where *p*_*t*_ is the predicted probability of the target class, *α* is the weighting factor for class imbalance, and *γ* is the focusing parameter that adjusts the rate at which easy samples are downweighted. Finally, to further improve the model’s ability to better discriminate between lacunes and mimics, we incorporated a pixel-wise supervised contrastive loss (*L*_*sup*_). The contrastive learning approach aims to bring the pixel embeddings of the same semantic class (lacunes) closer together while pushing apart the embeddings of different classes (lacunes vs. mimics/background). Therefore, we added a projection head after the penultimate layer of the *U*^2^Net decoder and applied pixel-wise supervised contrastive loss (Khosla et al., 2020) (Eqn. 3) to the embeddings produced by this projection head.

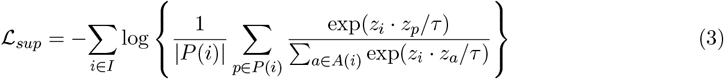

where 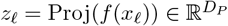 is the projected latent vector corresponding to the pixel *x*_*ℓ*_, *τ* ∈ ℛ^+^ is a scalar temperature parameter, *I* represents indices of all sampled pixels in the batch, the index *i* is the current *anchor, A*(*i*) represents all the indices except *i*, out of which *P* (*i*) represents the indices of positive pixels distinct from *i*, the · denotes the inner (dot) product and |*P* (*i*)| represents the cardinality of *P* (*i*). During training, for the first *N* epochs, negative pixels were randomly sampled from non-lacune pixels within the brain mask. After *N* epochs, when the model sufficiently learned the patterns, negative pixels were sampled from the false positives of the previous epoch. This approach motivated the model to focus on reducing false positives while improving the recall value. The total loss function for training the CPG module is given by,

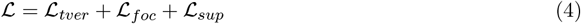

#### 2.2.1 Synthetic lesion simulation for robust training

The diverse appearance-based characteristics of lacunes, coupled with variations in scan parameters across multi-centre datasets (slice thicknesses ranging from 0.68 to 6 mm) and limited data, affected the sensitivity of the CPG module. To overcome this challenge, in addition to the composite loss function, we augmented our training data with simulated lacunes produced by a Synthetic Lesion Simulator (SLS).

The pipeline used to simulate lacune-like lesions using SLS is shown in Figure 3. In SLS, synthetic lesions were first generated on lacune-free slices of the dataset using the method proposed in our previous work, MedLesSynth-LD (Narayanan and Sundaresan, 2024). In line with emulating the shape and contrast variations of various lesions in MedLesSynth-LD, we created artificial lesions that emulated the structural, shape, and contrast characteristics of the lacunes in the simulation process. Synthetic lacunes were generated as localised structural perturbations of healthy brain tissue and constrained within randomly generated shape priors (formed using a set of ellipsoids). To preserve the inherent noise characteristics in the perturbed lesion textures in MRI, we used the Rician distribution (the underlying noise model for MRI (Narayanan and Sundaresan, 2024)) for the perturbation of healthy tissue. Next, to create a shape prior, we used a composite of ellipsoids to capture the shapes of both gliosis and cavitation in the lacunes. Finally, to match the sharp intensity difference observed between hyperintense gliosis and hypointense cavitation, we added perturbations within the two structures by varying the contrast parameters. These created lesions provided good priors, as they mimicked the structure, noise, and intensities of actual lesions. The computational overhead attached to MedLesSynth-LD is negligible because it does not require the training of deep learning models. To render the synthetic lesion much closer to the real lacunes (owing to their small size and variability between datasets), we translated the simulated lesions using cycleGAN (Zhu et al., 2017). For training the cycleGAN, patches of size 32 × 32 mm centred on the 2D real lacune instances were extracted to create the training data. Given that the typical maximum diameter of lacune is 15 mm, the selected patch size ensured that the patch encompassed the lacune with a sufficient amount of background tissue. Extracting localised patches allowed for the inclusion of multiple slices from the same lacune instance, enabling the model to learn inter-slice variations more effectively. The real-world patches with an equal number of simulated lacune patches were used to train the cycleGAN. Once trained, cycleGAN was used to translate the simulated lesion patches, which were then blended back into the original images using Poisson blending within a circular binary mask with a radius of 8 mm (empirically determined using trial and error), as shown in Figure 3.

**Figure 3:**
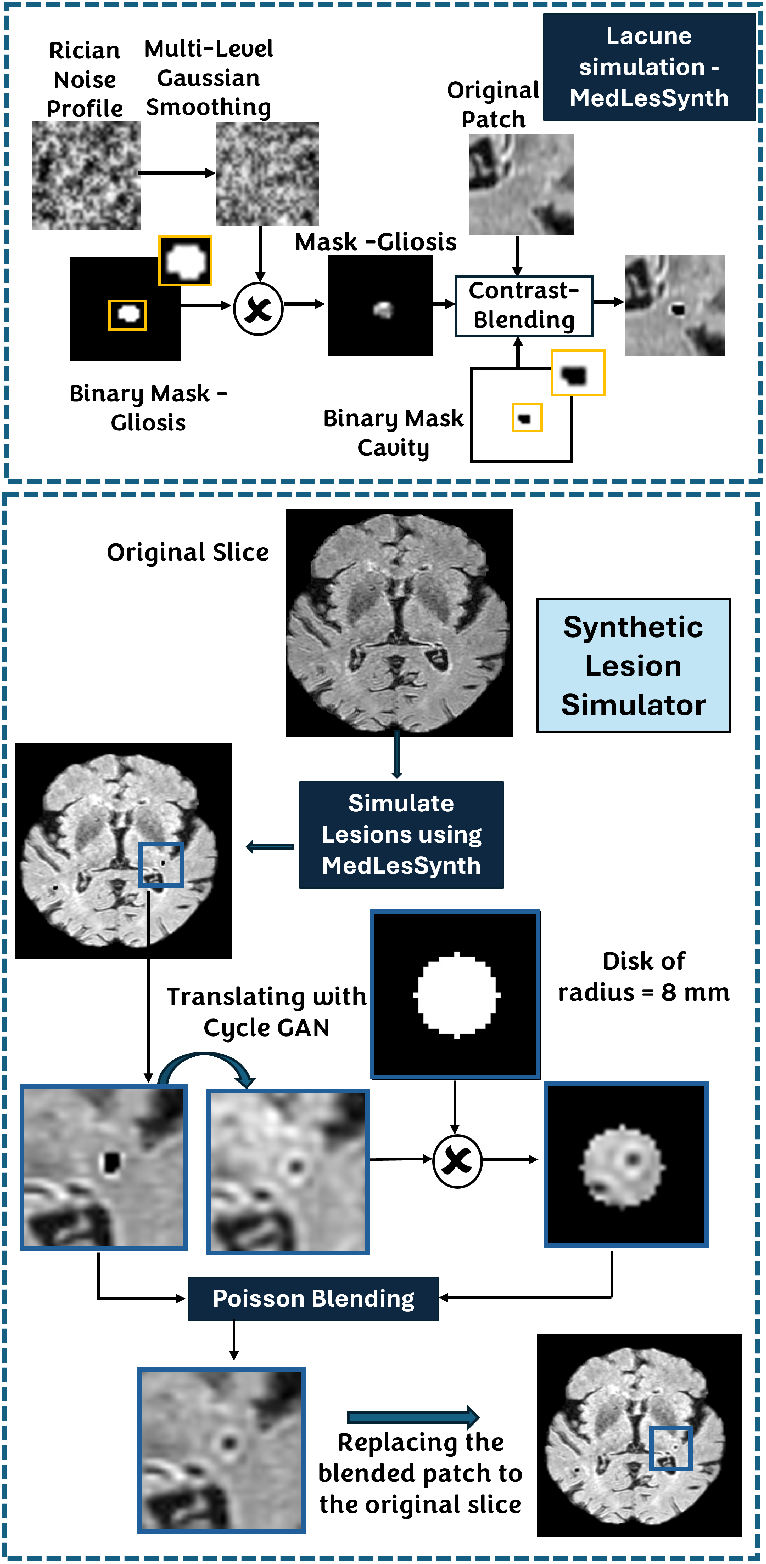
Lacune simulation pipeline using Synthetic Lesion Simulator (SLS). Top: Synthetic lacunes are generated using MedLesSynth-LD (Narayanan and Sundaresan, 2024), where healthy tissue is perturbed using a Rician noise model, within synthesised gliosis and cavity shape masks and applied on images at varying contrasts. Bottom: The synthesised lacunes are translated using cycleGAN and blended using Poisson blending to emulate the real lacune characteristics specific to the dataset.

### 2.3 SAM based Detection

As we aimed for high sensitivity in the CPG module, this typically resulted in false positives, mainly from parts of the sulcus and/or mimics such as EPVS, which were indistinguishable from the lacunes on the axial slices. Thus, the prompts generated from the CPG module were provided to the SAM module to reject false positives from slices at the volume level. We used the SAM model with the VIT-H encoder (Kirillov et al., 2023) on the entire 3D volume guided by point prompts from the CPG. This step essentially replicated the clinician’s approach of examining all three axes to distinguish lacune-like false detections from true lacunes.

Before applying SAM, we rescaled the intensities of the input volume using square-root transformation (for contrast enhancement) and performed min-max normalisation to facilitate better segmentation with SAM. The predicted 2D probability maps from the CPG module were then stacked together to obtain a 3D volume and thresholded at 0.5, to obtain 3D candidate instances. A threshold value of 0.5 was chosen experimentally based on the Free-Response Receiver Operating Characteristic (FROC) curve (shown in Figure 6). For each 3D candidate, its centroid was considered as the point prompt for SAM, and the axial, sagittal, and coronal slices (traversing through the centroid) were fed into SAM with the corresponding 2D coordinates as point prompts. Based on the segmentation output of the SAM, we detected true lacunes from the mimics, as explained below. Figure 4 shows a few examples of CPG-guided SAM-based detection of lacunes and the rejection of false positives.

**Figure 4:**
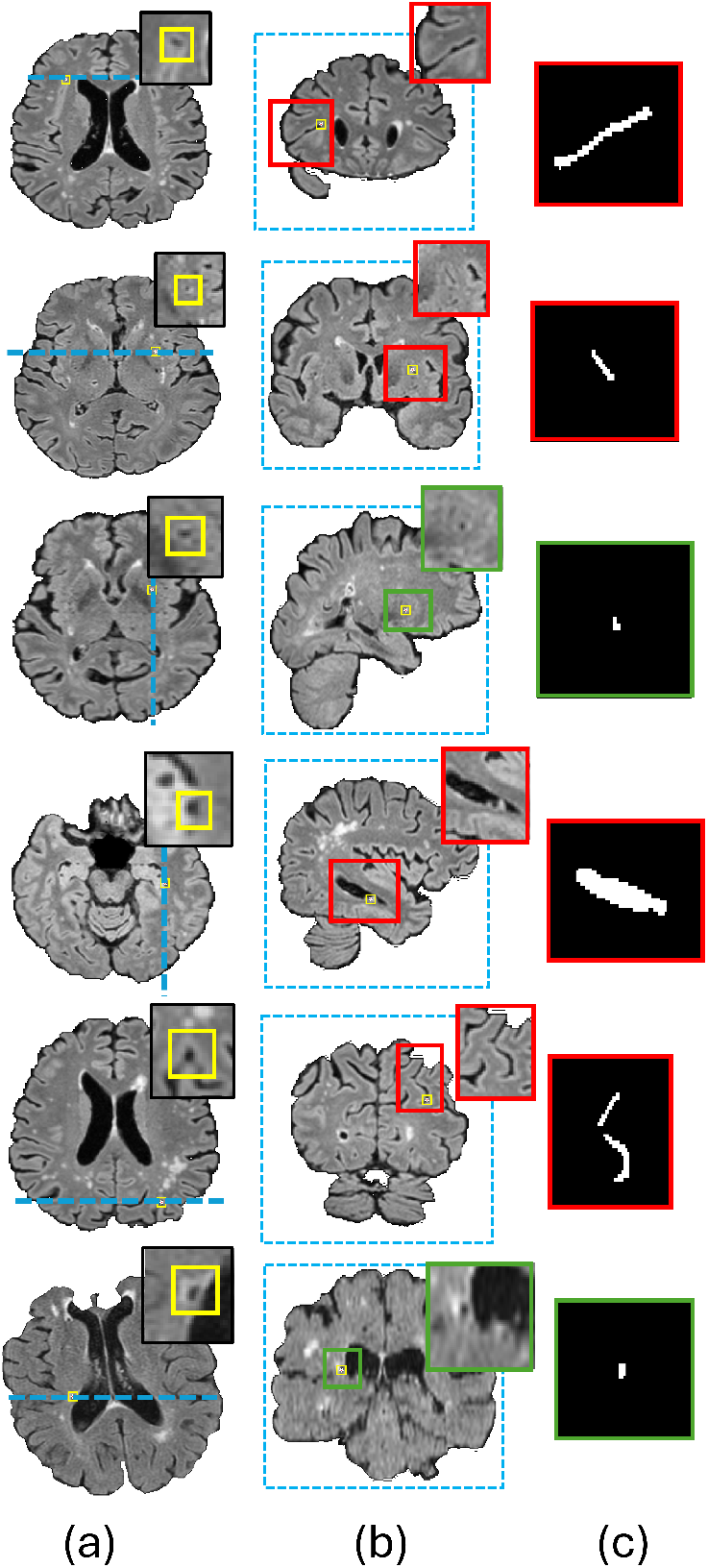
Lacune detection using SAM with point prompts from the Candidate Prompt Generator(CPG) module. Left to right: (a) candidates predicted by the CPG module (indicated by yellow boxes) shown on the axial slice, (also shown magnified at the top-right corner); the blue dotted line indicates the sagittal/coronal slice passing through the centroid, (b) the corresponding sagittal/coronal slice shown, with the magnified region of interest in the top-right corner, and (c) SAM’s predicted segmentation of the magnified region, with green and red boxes indicating the candidates predicted as lacunes and false-positives respectively.

- **Removal of sulci:** We removed the candidates that had SAM’s prediction probability ≥0.75 along any of the three axes, and intersected with the brain boundary, as sulci.
- **Removal of EPVS and other elongated structures:** EPVS might resemble lacunes in the axial plane but are more elongated in other planes. Among the three planes, if a lesion predicted by SAM had a major axis to minor axis ratio greater than 5 over any of the planes, then the instance was removed as a false positive. Despite the revised STRIVE criteria (Duering et al., 2023) allowing lacunes smaller than 3 mm, we eliminated SAM predictions with a maximum axial diameter of less than 3 mm to avoid misclassification of EPVS as lacunes. This is because the EPVS diameter typically lies within 3 mm, whereas we observed very few lacunes smaller than 3 mm.
- **Removal of false positives due to low tissue contrast:** SAM predictions with the confidence score *>*0.75 and predicted pixel area greater than 350 mm^2^ along any of the three axes were treated as false positives (due to segmentation of background tissue regions with low contrast) and eliminated. We determined the above threshold values of the confidence scores used in SAM empirically based on the trial-and-error method.

Furthermore, to account for the variation in slice thickness in the dataset, we adapted the following approach: for the slice thickness ≤ 4 mm, all three planes were inspected using SAM; for *>*4 mm, only the axial plane was examined using SAM, as a higher slice thickness led to less reliable structural integrity and continuity across slices. Note that this SAM-based methodology replicates clinical decision-making in distinguishing lacunes from mimics, leveraging both the 2D deep learning model’s initial prompts and SAM’s detailed segmentations in multiple planes to improve the overall lacune detection accuracy.

### 2.4 Adaptive region-aware thresholding

Lacunes have been shown to occur predominantly in specific regions of the brain, particularly in the white matter and subcortical structures (Duering et al., 2013; Benjamin et al., 2014). Hence, as the final step of our method, we utilised this location-based prior and used adaptive region-aware thresholding (ART) to further effectively reduce false positives. In this step, instead of applying a global threshold on candidates retained by SAM, we applied a region-wise threshold determined based on the prevalence of lacunes in different regions of the brain. Candidates were classified as false positives if the prediction softmax probabilities at the centroid of the candidates were below the ART thresholds for that region (shown in Figure 5). This adaptive threshold provided a more robust segmentation compared to applying a global threshold by providing a better trade-off in various regions. Additionally, it helped remove false positives that could not be filtered based on their attributes, as described in Section 2.3.

**Figure 5:**
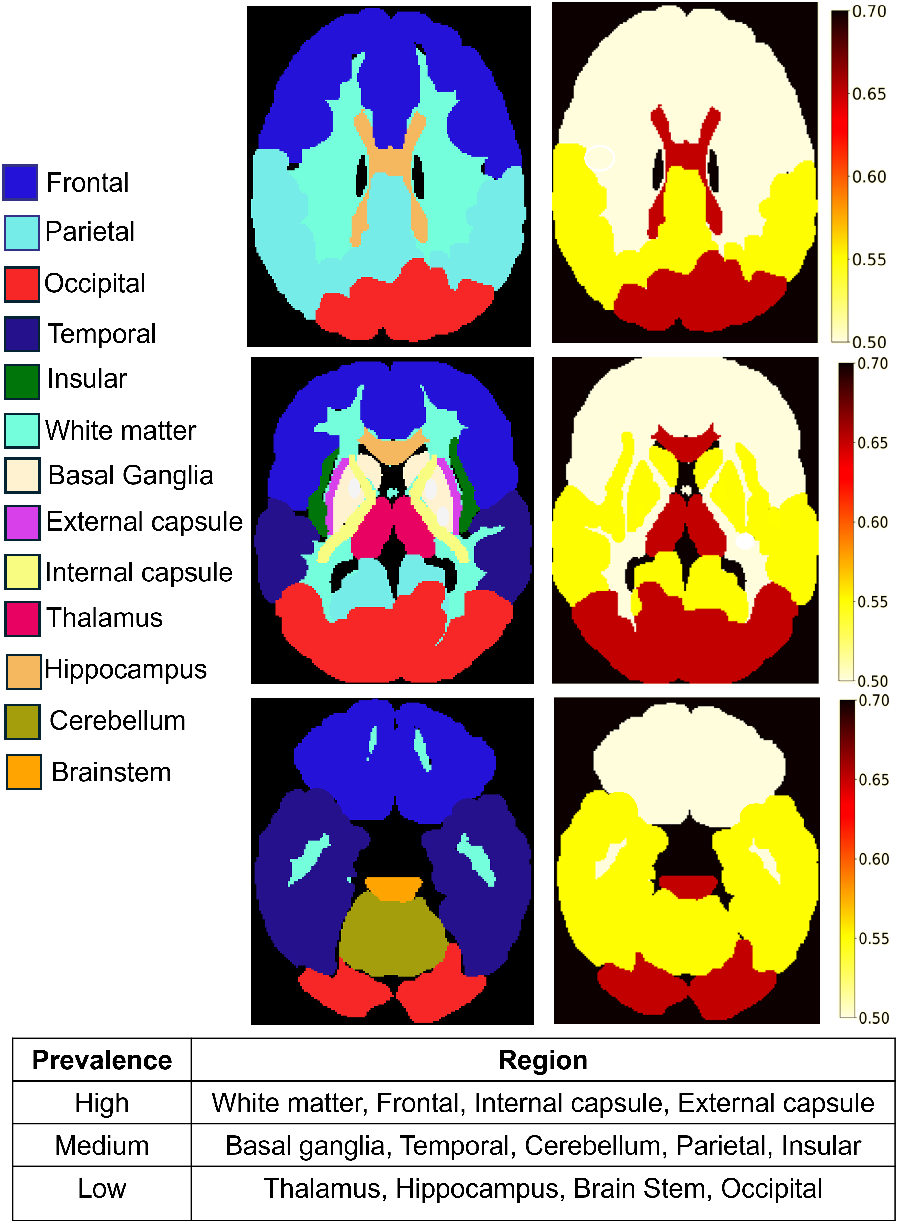
Parcellation of regions based on MARS scale for adaptive region-based thresholding. Top to bottom: Parcellation of regions according to the MARS atlas shown on the left with the corresponding threshold maps on the right for three different slices of the brain. The table shown below stratifies the brain regions based on the prevalence of lacunes for ART.

**Figure 6:**
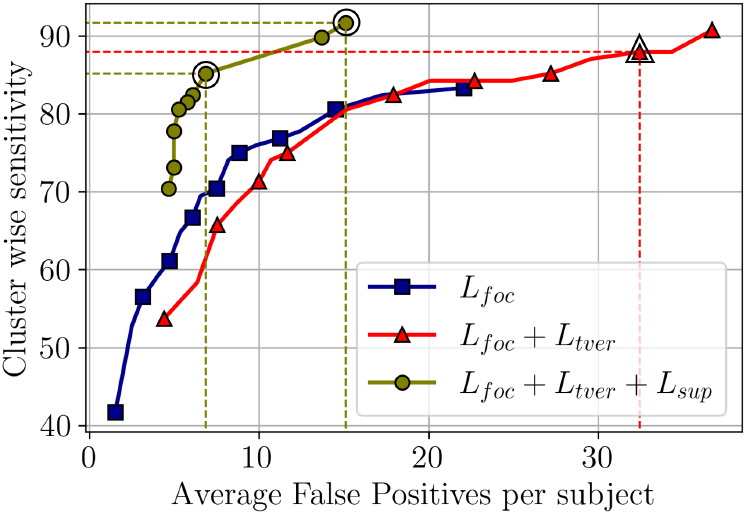
Ablation study results with different loss function components on the ISLES dataset. FROC curves illustrating CPG module performance with different components of loss functions, *L*_*foc*_, *L*_*foc*_ + *L*_*tver*_ and *L*_*foc*_ + *L*_*tver*_ + *L*_*sup*_, shown in blue, red and green curves respectively for the ISLES Dataset (the operating points that provide the best performance are indicated by dotted lines).

To analyse the region-wise distribution of lacunes in the dataset, we obtained segmentation of anatomical structures in the brain based on the microbleed anatomical rating scale (MARS) map (Gregoire et al., 2009). We chose the MARS map because it provided good granularity of the structural parcellation required for lacunes. The generation of MARS maps from various structural atlases in the MNI space was performed as specified by Sundaresan *et al*. (Sundaresan et al., 2025). Figure 5 shows the MARS atlas for three axial slices across the brain for a sample subject in MNI space. We registered the MARS atlas from the MNI space to the target subject space using the FSL FLIRT (Jenkinson et al., 2012). Once the anatomical regions were obtained in the subject space, we estimated the region-wise prevalence of lacunes in the training data by determining the lacune count in different regions. Finally, region-wise thresholds were assigned in alignment with the region-wise prevalence of lacunes (stratified as high/medium/low prevalence, as shown in Figure 5). The threshold values were 0.5, 0.55 and 0.65 for high, medium, and low values, respectively (note that these threshold values provided the best performance in the regions). The regions with a high prevalence of lacunes (specified ‘high’ in Figure 5) had lower threshold values compared to the regions with fewer lacunes (indicated as ‘low’ in Figure 5). Specifically, in the basal ganglia region where EPVS are usually more common (Wardlaw et al., 2013), a higher threshold (0.55) facilitated the removal of EPVS from predicted lacunes. Given that EPVS could closely mimic the appearance of lacunes, setting a higher threshold helped us retain more confident lacune predictions, while reducing low-confidence false positives representing EPVS.

### 2.5 Implementation Details

We implemented the proposed method using Python 3.11.7 and PyTorch 2.2.1. We trained the CPG module on a Linux workstation with an i9-10980XE CPU, 128 GB RAM, and an NVIDIA RTX A6000 GPU card with 48 GB memory. We used data augmentation techniques, including random rotation (*θ* ∈ [−10, 10] degrees) and horizontal and vertical flips. The training dataset for the CPG module contained 2,728 slices, including all positive slices and a subset of sampled negative slices from the ISLES dataset, along with 12,000 synthetic lacune slices generated using the SLS module (Section 2.2.1). The CPG module was trained using 14,728 2D slices with a 90%/10% training/validation split, requiring ≈6 min per epoch. We used the Adam optimiser with a learning rate of 1 × 10^−4^ and batch size of 8 for training. Regarding the loss functions in the CPG module, for *L*_*foc*_, we chose *α* = 0.25 and *γ* = 2 (as reported in (Ross and Dollár, 2017)). For *L*_*tver*_, we chose *β* = 0.97 and *α* = 0.03 to enhance the sensitivity of the model to the true lacune features. In our pixel-wise contrastive learning approach, for the initial *N* = 50 epochs, we sampled negative pixels from the background, whereas after 50 epochs, we sampled negative pixels from the previous epoch’s false positives. In the SLS module, we adopted the recommended loss functions and parameters for cycleGAN as mentioned in (Zhu et al., 2017). Unless specified, the above values were determined empirically using the trial-and-error method.

### 2.6 Dataset details

We used the following datasets for our experiments. As FLAIR is the only sequence common to both datasets, it was used exclusively in this study.

#### Ischemic Stroke Lesion Segmentation (ISLES) 2022 challenge training dataset (Hernandez Petzsche et al., 2022)

The primary dataset used in our study was the ISLES dataset, comprising of 250 MRI volumes, including FLAIR, DWI sequences, and ADC maps, collected from individuals aged 18 and older with diagnosed or suspected stroke. Data were collected from two different centres using 3T Philips, 3T Siemens, and 1.5T Siemens MAGNETOM MRI scanners. The slice thickness varied between 0.68 mm and 9.60 mm, as the dataset was originally curated to detect larger lesions such as strokes. However, for lacune detection, we excluded volumes with slice thicknesses greater than 6 mm and those with heavy artefacts, as these are not suitable for detecting smaller lesions such as lacunes. Lacune annotations were performed with the assistance of an expert radiologist using the ITK snap (Yushkevich et al., 2006) (a free, open-source software application developed at the University of North Carolina). This resulted in a final set of 221 volumes consisting of 1,021 lacunes, of which 181 volumes were used for training, validation and 40 volumes for testing.

#### VAscular Lesions Detection and segmentatiOn challenge 2021 (VALDO) dataset (Task-3)

This dataset, available as VALDO task-3 training data, comprised MRI data from 40 patients from two different cohorts: 6 from the Southall and Brent Revisited (SABRE) (Tillin et al., 2012; Jones et al., 2020) and 34 from the Rotterdam Scan Study (RSS) (Ikram et al., 2015). The RSS cohort included participants aged ≥ 45 years who underwent follow-up every 3-4 years to investigate chronic illness. In SABRE, data were collected from individuals aged 36–92 years (mean age: 72 years) to monitor metabolic and cardiovascular diseases. The imaging protocol included T1-weighted, T2-weighted, and FLAIR sequences acquired using either a 3T Philips (SABRE) or 1.5T GE (RSS) scanner. Thus, the image resolutions varied between cohorts: SABRE had a resolution of 1.09 × 1.09 × 1.09, while RSS had 0.49 × 0.49 × 0.8 mm for all sequences. Manual annotations of lacunes by two independent raters were available for each volume of this dataset. The dataset consisted of 83 lacunes. We used 20 volumes for training and validation and 20 volumes for testing.

## 3 Experiments

### 3.1 Evaluation of various model architectures for candidate detection

In the CPG module, for detecting lacune candidates to generate prompts for SAM, we evaluated different model architectures, including *U*^2^Net (Qin et al., 2020), UNet++ (Zhou et al., 2018), ResUNet++ (Jha et al., 2019), Attention UNet (Oktay, 2018), and Swin UNETR (Hatamizadeh et al., 2021). These architectures were trained using the methodology outlined in Section 2.2 with the parameters specified in Section 2.5 on the ISLES data, and evaluated using the metrics specified in Section 3.7.

### 3.2 Ablation Study I: Effect of components of the loss function on the CPG performance

We performed an ablation study to investigate the effect of adding different loss components (Eqn. 4) on the candidate detection performance of the CPG module. For this purpose, we compared the performance of the CPG module by incrementally adding the components of the loss function as follows: (1) *L*_*foc*_, (2) *L*_*foc*_ + *L*_*tver*_, and (3) *L*_*foc*_ + *L*_*tver*_ + *L*_*sup*_. We evaluated performance on the ISLES dataset using the metrics described in Section 3.7.

### 3.3 Ablation study II: Effect of different modules on the proposed lacune detection method

We also conducted an ablation study to examine the impact of various modules in the lacune detection workflow on the ISLES dataset. For this, we incrementally integrated the modules as follows: (1) candidate detection using CPG, (2) detection of lacune candidates by SAM using prompts generated by CPG (CPG+SAM) by applying a global threshold of 0.5, and (3) applying the adaptive thresholds using ART on SAM output (CPG+SAM+ART). We evaluated the performance enhancement with successive integration of modules using the metrics specified in Section 3.7.

### 3.4 Effect of using simulated lesions from SLS on the CPG performance

The ISLES dataset was multi-centric data (with two different centres) with different slice thicknesses and lacunes with varying appearances and features. Hence, to address the challenge posed by the limited training data in learning the diverse characteristics of lacunes, we used simulated lesions generated by the SLS module (described in Section 2.2.1) to expand the training set. We evaluated the effect of augmenting the training data with simulated lesions on the performance of the CPG module for the ISLES dataset using the metrics specified in Section 3.7. Additionally, we plotted t-distributed stochastic neighbour embedding (t-SNE) plots and determined the Frechet inception distance (FID) scores (Heusel et al., 2017) to demonstrate the similarity between the simulated and real lacunes. Note that the lower the FIR scores between the simulated and real lacunes, the better is their similarity.

### 3.5 Self-Distillation for robust lacune candidate detection in a low-data regime

The second dataset in this study, the VALDO dataset, contained only a few training samples and exhibited a domain shift, as the data were obtained from two different cohorts. To achieve good performance in this low-data case for the CPG module, it was crucial for the model to retain the knowledge about lacune characteristics learned from a larger dataset, such as ISLES, while adapting to the lesion characteristics within the VALDO dataset. Hence, we implemented self-distillation as a strategy to adapt our method for generalisation across datasets. Self-distillation has been proven to be effective in prior studies (Paluru et al., 2023; Kim et al., 2020; Kuang et al., 2024), where the model uses its own predictions from a previous epoch as additional training input for subsequent epochs. In this approach, we pretrained the model on the ISLES dataset for the initial 100 epochs. Then, each subsequent epoch was trained as follows: In the first step, the model was trained on the ISLES dataset, with its current state acting as a *teacher* for the next step within that epoch. In the next step, the same model was trained on the smaller VALDO dataset using the lacune ground truth as hard labels and the teacher’s predictions as soft labels, with a distillation loss described in Eqn. 5 with *α* = 0.65. We experimented with various values of *α* and observed that *α* = 0.65 was optimal. Furthermore, because the predictions for VALDO were made using the self-distilled model, which produces more confident predictions, the optimal ART thresholds for the VALDO dataset were incremented by a factor of 0.1.

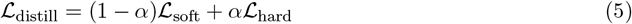

where ℒ_hard_ represents the Focal loss (ℒ_*foc*_, described in eqn. 2), and ℒ_soft_ represents the KL divergence between the student and teacher logits.

We evaluated the performance of the CPG module on the VALDO dataset using the metrics specified in Section 3.7 with different training strategies, including combinations of both datasets and self-distillation.

### 3.6 Comparison with existing methods

Finally, we compared our performance with that of existing state-of-the-art methods for lacune detection. We included deep learning-based semi-automated and fully automated methods for lacune detection proposed since 2017 in our comparison.

### 3.7 Performance evaluation metrics and statistical evaluation

We used the following evaluation metrics consistent with existing methods (Ghafoorian et al., 2017; Al-Masni et al., 2021; Sudre et al., 2024): (1) *cluster-wise sensitivity*, which is the ratio of the number of true positive lacune lesions detected by the model to the total number of true lacune lesions across the entire test set, (2) *cluster-wise F1-measure*, which is the harmonic mean of cluster-wise precision and cluster-wise sensitivity, and (3) *Absolute Element Difference (AED)*, the absolute difference between the number of true lacune lesions and the number of predicted lacune lesions.

Because the VALDO dataset includes annotations from two raters, we determined the F1-score weighted by inter-rater agreement and average AED, adhering to the metrics used in the VALDO 2021 challenge (Sudre et al., 2024). For statistical evaluations, we used a Wilcoxon signed rank test to assess the statistical significance of performance differences (p*<*0.05 for significance; p*<* 0.001 for stronger significance) in the ablation studies.

## 4 Results

### 4.1 Evaluation of different model architectures for Lacune Detection

The results of the comparison of the various model architectures on the ISLES dataset are presented in Table 1. Among the architectures evaluated, Attention UNet achieved the best F1-score and AED. However, we selected *U*^2^Net for the CPG module because the primary focus of the module was to maximise the sensitivity for the detection of true lacunes. At a threshold of 0.5, *U*^2^Net achieved the highest sensitivity of 85.19%, thus making it the optimal choice.

**Table 1:** Comparison of performance of different deep learning models for CPG module for the ISLES Dataset, with number of parameters and FLOPS (in Giga(G) FLOPS). (↑)/(↓) indicates that higher/lower values are better. The best sensitivity value is reported in bold. The median values (with the interquartile range in brackets) are reported for F1 value and AED.

| Model | Number of Parameters | Number of FLOPS | F1 value (↑) | AED (↓) | Sensitivity (↑) |
| --- | --- | --- | --- | --- | --- |
| $U^2$Net | 1.17M | 12.63G | 45.30[23.75 – 63.23] | 5[2.75 – 8.25] | **85.19** |
| Attention UNet | 34.91M | 68.82G | 49.90[18.83 – 72.50] | 2[1 – 11] | 78.71 |
| Res UNet++ | 4.06M | 15.68G | 31.14[13.98 – 66.67] | 3.5[1 – 14.5] | 71.30 |
| Unet++ | 2.42M | 18.31G | 34.13[20.24 – 51.79] | 4[2.75 – 8.25] | 62.04 |
| Swin UnetR | 6.31M | 4.9G | 27.85[16.41 – 49.23] | 6[3 – 12.50] | 60.18 |

### 4.2 Ablation Study I: Effect of components of the loss function on the CPG performance

Figure 6 shows the FROC curves indicating the effect of the addition of different loss components on the CPG module for the ISLES dataset. Although focal loss aids in learning more difficult examples, the maximum sensitivity achieved was 84%. With the addition of the Tversky loss with a higher weight of 0.97 for false negatives, the model captures more true lacunes, providing a sensitivity of around 90%, despite greater heterogeneity in the appearance of the lesion. However, the average number of FPs was 32/volume, which would incur higher computational costs in the subsequent stages. With the inclusion of the contrastive learning component in the loss function, the model learned tissue characteristics more effectively, providing far fewer FPs of 15/volume and 6.9/volume for sensitivities of 91% and 85.19%, respectively (as highlighted in Figure 6). For subsequent analysis, we selected a high-precision operating point with an 85.19% sensitivity.

### 4.3 Ablation study II: Effect of different modules on the proposed lacune detection method

The ablation study results are presented in Table 2 and the visual results are shown in Figure 7. As shown in Table 2, the integration of each module led to an improvement in the F1-scores for both the ISLES and VALDO datasets. The improvements were statistically significant in almost all cases (except for the addition of ART to the VALDO dataset). While the CPG module demonstrated strong performance in capturing true lacunes, the SAM module proved to be very effective in reducing false positives originating from structures such as the sulci, cerebral ventricles, EPVS, and other regions, by analysing candidates along the three axes. In this process, the SAM module retained all the true lacunes and markedly improved the metrics across both datasets. Although the SAM module offered substantial improvements in metrics across both datasets, its ability was constrained by the heterogeneity in the appearance of lacunes. Integration of the ART module helped overcome this constraint by using adaptive thresholds based on knowledge of the region-wise distribution of lacunes. This further improved detection accuracy by eliminating false positives that closely resembled lacunes on all axes, as shown in Figure 7. Consequently, the proposed method achieved an overall sensitivity of 84.26% for the ISLES dataset and 92.31% for the VALDO dataset.

**Table 2:** Ablation Study: Performance metric with the addition of different modules. CPG, SAM, ART represents Candidate prompt Generator, SAM based lacune detection and Adaptive Region-aware thresholding respectively. (↑)/(↓) indicates that higher/lower values are better. The median values (with the interquartile range in brackets) are reported for F1 value and AED. */** indicates p*<*0.05/p*<*0.001 showing significant difference between successive rows for F1 value and AED.

| Module | F1 value (↑) | AED (↓) | Sensitivity (↑) |
| --- | --- | --- | --- |
| ISLES Dataset |  |  |  |
| CPG | 45.30 [23.75-63.23] | 5.00 [2.75-8.25] | 85.19 |
| + SAM | 57.14 [27.68 – 79.89]** | 3.00 [1 – 6]** | 85.19 |
| + ART | 67.38 [37.14 – 82.50]* | 2.00 [1-5] | 84.26 |
| VALDO Dataset |  |  |  |
| CPG | 44.44 [32.22 – 54.07] | 7.75 [2.75 – 11.25] | 92.31 |
| + SAM | 66.67 [50.00 – 70.24]* | 2.25 [1 – 4.25]** | 92.31 |
| + ART | 69.92 [51.96 – 74.11] | 1.50 [1 – 3]* | 92.31 |

**Figure 7:**
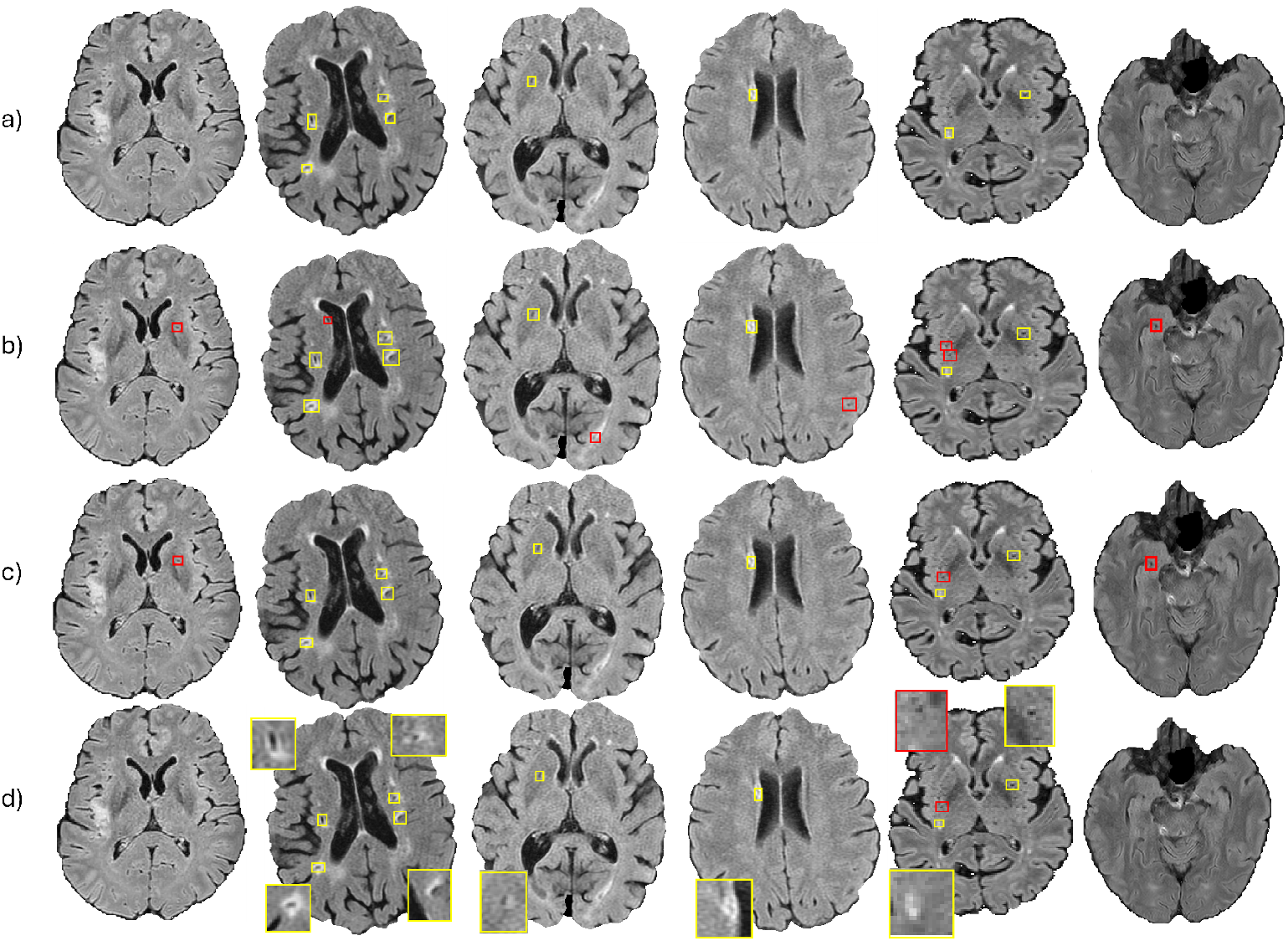
Sample results of the proposed method from the ISLES and VALDO datasets. Top to bottom: a) axial slice with ground truth (shown in yellow boxes), b) prompts generated by the CPG module, c) predictions from SAM module to reduce false positives and d) detected final lacunes from Adaptive Region-aware Thresholding (ART). In (b)-(d), the yellow and red bounding boxes indicate the true lacunes and false positives respectively.

### 4.4 Effect of using simulated lesions from SLS on the CPG performance

The t-SNE plot in Figure 8 (top panel) illustrates the similarity between the real and simulated lacunes by showing good overlap between them. Hence, the lacunes generated by our SLS module closely resembled the characteristics of real lacunes while effectively capturing the heterogeneity observed in the actual dataset. Notably, despite the small size of the lacunes and wide variability in their appearance, the lacunes simulated using MedLesSynth and those further refined through cycleGAN achieved promising FID scores of 68.36 and 33.56, respectively, with respect to real lacunes. Furthermore, the impact of incorporating simulated lesions during training is evident from the FROC curve shown in Figure 8 (bottom panel). When the CPG module was trained without simulated lesions, using only 2,724 slices from the ISLES dataset (as indicated in Section 2.5), it achieved a sensitivity of 79%, despite allowing an average of 21 FPs/subject. In contrast, the inclusion of simulated lesions expanded the training set to 14,724 slices, enabling the CPG module to achieve a higher sensitivity of 91% while also reducing the average FPs to 15/subject, highlighting the effectiveness of the simulated lesion in enhancing the performance.

**Figure 8:**
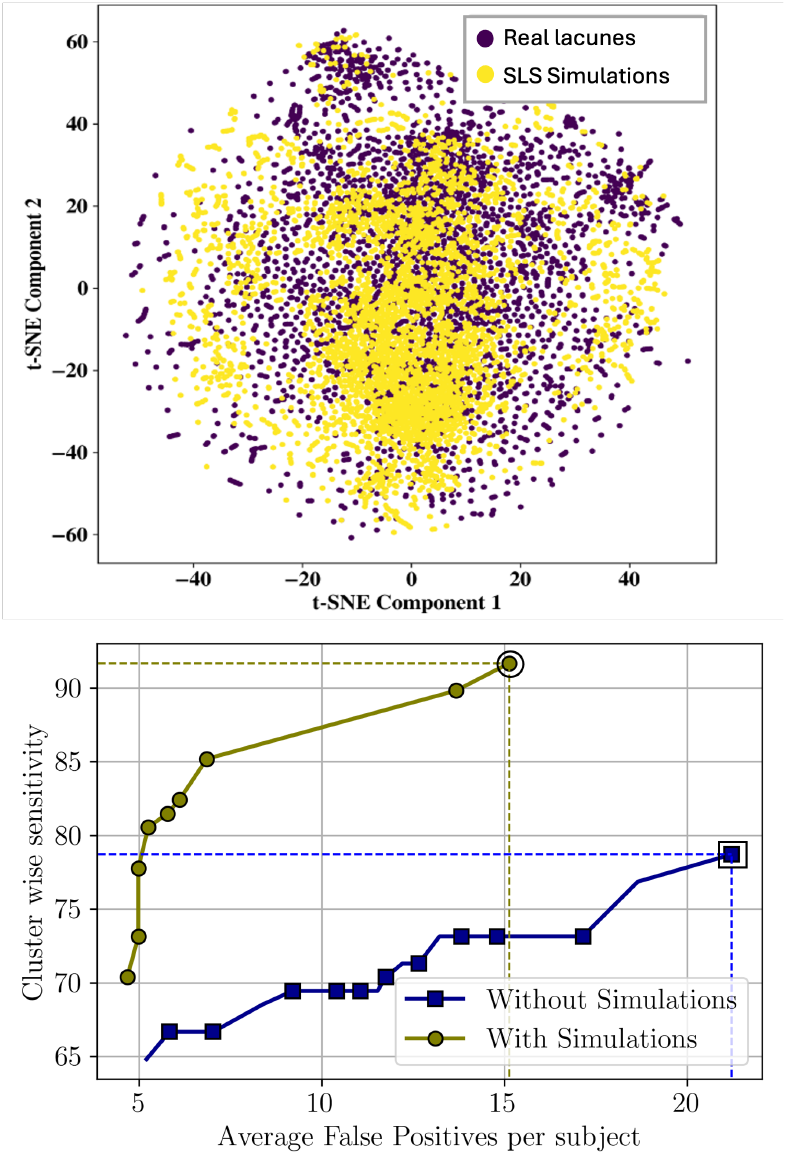
Effect of simulations from SLS module on CPG performance. Top: T-SNE plot showing good oevrlap between real-world lacunes and synthetic lacunes simulated using SLS. Bottom: FROC curves showing the comparison of the performance of the CPG module trained with and without inclusion of simulated data (green and blue curves respectively) for the ISLES Dataset (the operating points that provide the best performance are indicated by dotted lines).

### 4.5 Self-Distillation for robust lacune candidate detection in a low-data regime

Table 3 reports the results of the comparison of CPG detection performance for self-distillation and with training on a combined dataset (VALDO + ISLES) on the VALDO dataset. The self-distillation approach significantly improved the lacune detection performance of the CPG module, achieving a sensitivity of 92.1% for lacune candidates annotated by both raters. The ISLES dataset was acquired from individuals with suspected or diagnosed stroke, whereas the VALDO dataset was collected from an asymptomatic population. This resulted in significant variations in the appearance of lacunes, which were influenced by differences in lesion location and age. Owing to these disparities in lacune characteristics, the model trained on the combined dataset could not adapt exclusively to the VALDO dataset, and hence showed reduced sensitivity. This was further exacerbated by a significant sample size imbalance between the VALDO and ISLES datasets. However, self-distillation effectively captured true lacunes in the VALDO dataset, despite a slight increase in AED, which was addressed in subsequent modules to improve the performance.

**Table 3:**
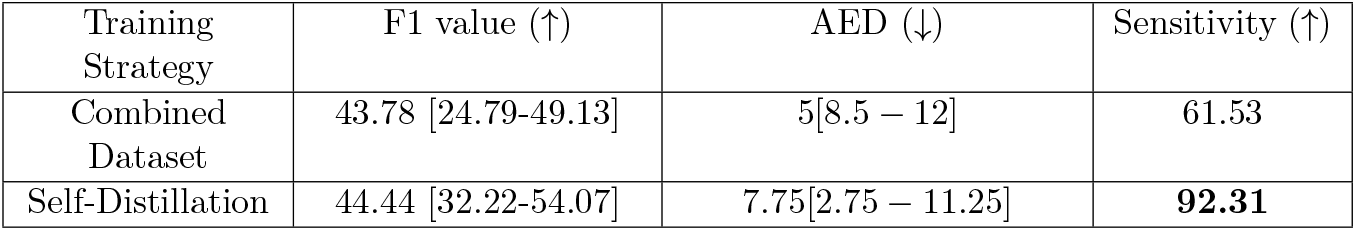
Comparison of the CPG module’s performance on the VALDO dataset using a combination of both datasets and self-distillation as training strategies, with the best sensitivity value reported in bold. (↑)/(↓) indicates that higher/lower values are better. The median values (with the interquartile range in brackets) are reported for F1 value and AED.

### 4.6 Comparison with existing methods

The proposed lacune detection method demonstrated superior performance compared with existing approaches across multiple metrics, as shown in Table 4. It consistently achieved high accuracy across varying image resolutions, with significantly lower false positive rates of 0.05 and 0.06 per slice for both datasets which is less than half the rate reported by Ghafoorian et al. (Ghafoorian et al., 2017). This significant reduction in false positives can be attributed to our novel modular strategy of examining candidates across all three axes using SAM and integrating knowledge of the regional distribution of lacunes.

**Table 4:** Comparison of the performance of proposed method for detection of lacunes with the existing semi-automated and automated methods. (↑)/(↓) indicates that higher/lower values are better. The median values are reported for F1 value and AED.

Semi-Automated methods
| Method | Sequence [Slice Thickness] | No of Lacune Instances | Sensitivity (↑) | Average FPs (↓) | Remark |
| --- | --- | --- | --- | --- | --- |
| Al-Masni et al.(2021) | T1 [1mm], FLAIR [2mm] Uniform Resolution | 696 | 96.41 | 1.32 /subject | Manual candidate detection followed by automated candidate discrimination. |

Automated methods
| Method | Sequence [Slice Thickness] | No of Volumes [No of Lacune Instances] | Sensitivity (↑) | Average FPs (↓) | F1 value (↑) | AED (↓) |
| --- | --- | --- | --- | --- | --- | --- |
| Ghafoorian et al.(2017) | T1 [1mm], FLAIR [3mm] Uniform resolution | 111 [38] | 97.4 | 0.13 /slice | - | - |
| Sudre et al.(2019) | T1 [1mm], T2[1mm], FLAIR[1mm] Uniform resolution | 2 | 72.7 | - | - | - |
| Team Tea.(2024) | T1 [0.8mm, 1mm], T2 [0.8mm, 1mm], FLAIR [0.8mm, 1mm] | 66 [185] | - | - | 28.57 | 1 |
| Ensemble Top-VALDO Challenge (2024) | T1 [0.8mm, 1mm], T2 [0.8mm, 1mm], FLAIR [0.8mm, 1mm] | 66 [185] | - | - | 30.77 | 1 |
| Proposed | VALDO - FLAIR [0.8mm, 1mm] | 20 [38] | 92.31 | 0.06 /slice | 69.92 | 1.5 |
| Proposed | ISLES - FLAIR [0.68mm to 6mm] | 40 [108] | 84.26 | 0.05 /slice | 67.38 | 2.00 |

Although Ghafoorian et al.’s approach achieved a sensitivity of 97.4% for lacunes agreed upon by three out of four raters, with 0.13 false positives per slice, their false positive rate was significantly higher than our reported rate of 0.05 per slice, corresponding to a sensitivity of 84.26% on lacunes agreed upon by any two of the four raters involved in their study. In addition, while their model was evaluated on uniform resolution volumes from a single scanner model, our approach demonstrated greater versatility, maintaining strong performance across multi-resolution data from four different imaging centres. In particular, our method demonstrated improved performance compared to the top entries in the VALDO challenge, achieving a sensitivity of 92.31% on lacune candidates agreed upon by both raters in VALDO and 84.26% in ISLES, supported by higher F1 scores computed for comparable numbers of lacunes across both datasets. The method proposed by Duan *et al*. (Duan et al., 2020) achieved a region-wise F1 score of 68.3, while our method achieved an on-par F1-score on a volume-wise basis, despite the fact that the region-wise F1 scores typically surpass volume-wise scores. Furthermore, unlike existing methods that depend on multiple imaging sequences for decision making (summarised in Table 4), our approach maintains optimal performance while relying solely on FLAIR sequences.

## 5 Discussion

In this study, we proposed a modular framework for lacune detection that integrates lacune candidate prompt generation, SAM-based morphological examination, and adaptive thresholding for false positive reduction. Given the wide range of appearance-based heterogeneity, inter-slice variations, and multi-centre datasets with limited training data from specific target datasets, the proposed method augmented the training data using simulated lesions and used contrastive learning and knowledge distillation approaches to ensure the accurate detection of lacunes. Using only FLAIR sequences, our method achieved sensitivities of 84.26% and 92.31% on the ISLES and VALDO datasets, respectively, with corresponding F1 scores of 67.38% and 69.92%, respectively, compared favourably with existing state-of-the-art approaches.

In general, the detection of lacunes is challenging owing to their small size and heterogeneous appearance, and often leads to diagnostic disagreement, even among experienced clinicians, as evidenced in previous studies (Ghafoorian et al., 2017; Sudre et al., 2024; Duan et al., 2020). These challenges resulted in existing approaches reporting a higher number of false positives, as shown in Table 4. The existing methods also exhibit limitations when processing datasets with varying resolutions and cannot efficiently handle volumes of higher slice thicknesses. This is particularly common in stroke imaging (given that lacunes are crucial biomarkers for stroke diagnosis), which typically involves larger slice thicknesses for quicker acquisition. Hence, an automated lacune detection method should be capable of working reliably across a range of image resolutions, irrespective of its small size. Our methodology successfully addressed these limitations by implementing specialised modules, each targeting distinct aspects of lacune detection. The initial CPG module identified potential lacune candidates with high sensitivity, although it allowed for some false positives. The CPG module used the U^2^Net model, which incorporated multiple Residual U-blocks (RSUs) that effectively processed features across various receptive fields while preserving fine details. This is a crucial requirement for detecting small lesions, such as lacunes, thus providing sufficiently high sensitivities of 85.19% and 92.31% for the ISLES and VALDO datasets, respectively, as indicated in Table 2. Furthermore, our data augmentation strategy implemented a lesion simulation process that generated realistic lacune lesions using shape- and noise-based texture priors, followed by translation using a cycle-GAN to match the dataset-specific lacune characteristics. This two-phase approach proved essential for lacune simulation, as in general, GANs struggle with the generation of small lesions from scratch, while simple contrast blending of shape and texture priors failed to accurately replicate actual lacune characteristics because of their small size and unique appearance (hypointense centre with a hyperintense rim). However, the inclusion of simulated lesions for training addressed the challenge of limited data, generating synthetic lesions with typical lacune characteristics with sufficient diversity. This enabled the model to learn the complex characteristics of lacunes better, leading to an improvement in the sensitivity of the CPG module from 75% to 91%, while significantly reducing false positives (as shown in Figure 8 (bottom panel)). Finally, the training process of the CPG module utilized 2D axial slices and employed a combination of focal, Tversky, and contrastive loss functions, enhancing the module’s capability to learn subtle inter-slice variations and challenging lacune examples while differentiating tissue characteristics (as shown in Figure 6).

In the proposed method, the SAM module played a major role in the reduction of false positives and effectively mimicked the clinical expert’s decision making for distinguishing true lacunes from the mimics. In the SAM-based selection module, we examined potential lacunes from all three anatomical axes to distinguish true lacunes from false positives, such as sulci, EPVS, and low-contrast regions. SAM proved particularly well suited for this task because of its zero-shot segmentation capabilities (Kirillov et al., 2023), offering precise segmentation of lacunes when provided with representative point prompts from candidates, regardless of domain shifts. We strategically extracted point prompts (rather than bounding boxes) to enable unrestricted segmentation along elongated structures such as sulci or ventricles, provided the prompt was positioned well within the structure. Additionally, the ART module helped eliminating challenging mimics that closely resembled lacunes (when viewed from all axes, thus being overlooked by SAM) by employing adaptive thresholds. For this, we utilised the MARS map which was originally designed as a standard region-based atlas for rating another CSVD sign: cerebral microbleeds. In addition to the common risk factors associated with both signs, the MARS map also provided detailed parcellation of the brain that was sufficient to analyse the spatial distribution of lacunes. To the best of our knowledge, this is the first approach to use population-level insights and lacune incidence to define region-specific adaptive thresholds, filter false positives, and improve the precision of lacune detection.

The datasets used in our evaluation were distinct with notably different characteristics. The ISLES dataset comprised patients with diagnosed or suspected stroke (from two different centres), whereas the VALDO dataset was collected from individuals without dementia who were being monitored for chronic illnesses, metabolic conditions, and cardiovascular diseases (from 2 different cohorts), as detailed in Section 2.6. These distinct patient populations exhibit markedly different lacune characteristics. For instance, lacunes in the ISLES dataset were predominantly extensively cavitated, with highly irregular, thin, or completely absent hyperintense rims. In contrast, the lacunes in the VALDO dataset featured prominent hyperintense rims and small cavitated areas, consistent with the asymptomatic population. For the VALDO dataset, despite the significant domain shift from the ISLES dataset, varying appearance characteristics, and limited size of the VALDO training set, our method achieved its highest performance owing to our self-distillation strategy for initial prompt generation, as indicated in Table 3. This approach focused on minimising distillation loss and aligning the student model’s outputs with the teacher’s softened predictions, which allowed the model to learn high-level abstractions that best discriminated lacunes from the mimics, which were invariant across datasets.

Our method compared favourably with the existing method and achieved high detection sensitivity with minimal false positives at rates of 0.05 and 0.06 per slice for the ISLES and VALDO datasets, respectively. However, there is still scope for improvement in the generalisability and computational complexity of the proposed method. For instance, the limitations include the additional computation required to train the cycleGAN model for synthetic lacune simulations on individual datasets to match its specific characteristics. In addition, the adaptive threshold values relied on the lacune features learned by the model on the individual datasets, hence segmenting lacunes with higher/lower softmax scores. Note that despite the minor increment in the threshold values, the overall trend of the threshold across different regions remained consistent (high *>* medium *>* low), thus illustrating the importance of adaptive thresholds.

## 6 Conclusions

The proposed method achieved sensitivity values of 84.26% and 92.31%, with average FP of 0.05 and 0.06 per slice, and median F1-scores of 67.38 and 69.92 on the ISLES and VALDO datasets, respectively, outperforming the existing state-of-the-art methods. The proposed method could enable the determination of lacune counts in various brain regions, which is crucial for analysing the spatial distribution of lacunes and their joint effect with other comorbidities related to neurodegeneration. Future research directions include studying the population-level correlation between lacune characteristics and several clinical/demographic factors, with a focus on risk prediction and prognosis for various neurological and neurodegenerative diseases.

The code files have been made available at https://github.com/PonDeepika/Lacune_Detection

## Data Availability

All data produced are available online at
1) https://zenodo.org/records/7153326
2) https://zenodo.org/records/4520773

https://zenodo.org/records/4520773

## CRediT authorship contribution statement

Pon Deepika: Conceptualization, Methodology, Software, Validation, Investigation, Formal analysis, Writing - original draft, Visualization. Gouri Shanker: Writing - review & editing, Validation, Investigation. Ramanujam Narayanan: Writing - review & editing, Validation, Investigation. Vaanathi Sundaresan: Conceptualization, Writing – review & editing, Supervision, Resources, Project administration, Funding acquisition, Data curation.

## Ethics Statement

For the VALDO dataset, all cohorts were retrospective studies for which local ethical approval had already been obtained from the National Research Ethics Service Committee, London-Fulham (14/LO/0108) for SABRE and the Population Research Act from the Ministry of Health for RSS. For all datasets, acquisition of the data was performed by a trained radiographer according to a predefined research protocol. For the ISLES dataset, the retrospective evaluation of imaging data was approved by the local ethics boards of all participating centers. Requirement of written informed consent was waived by all ethics boards due to the retrospective nature of the study and the rigorous patient de-identification of the data.

## Acknowledgements

The VALDO Dataset study was funded by Wellcome Trust (082464/Z/07/Z), British Heart Foundation (SP/07/001/ 23603, PG/08/103, PG/12/29/29497 and CS/13/1/30327), Erasmus MC University Medical Center, the Erasmus University Rotterdam, the Netherlands Organization for Scientific Research (NWO) Grant 918-46-615, the Netherlands Organization for Health Research and Development (ZonMW), the Research Institute for Disease in the Elderly (RIDE), and the European Union Seventh Framework Programme (FP7/2007–2013) under grant agreement No. 601055, VPHDARE@IT, the Dutch Technology Foundation STW.

## Data availability

The ISLES dataset is publicly available at https://zenodo.org/records/7153326

